# Seizure Onset Zone Identification Using Phase-Amplitude Coupling and Multiple Machine Learning Approaches for Interictal Electrocorticogram

**DOI:** 10.1101/2021.10.27.21265585

**Authors:** Yao Miao, Yasushi Iimura, Hidenori Sugano, Kosuke Fukumori, Toshihisa Tanaka

## Abstract

Automatic seizure onset zone (SOZ) localization using interictal electrocorticogram (ECoG) improves the diagnosis and treatment of patients with medically refractory epilepsy. This study aimed to investigate the characteristics of phase-amplitude coupling (PAC) extracted from interictal ECoG and the feasibility of PAC served as a promising biomarker for SOZ identification. We employed the mean vector length modulation index approach on the 20-s ECoG window to calculate PAC features between low-frequency rhythms (0.5–24 Hz) and high frequency oscillations (HFOs) (80–560 Hz). We used statistical measures to test the significant difference in PAC between SOZ and non-seizure onset zone (NSOZ). To overcome the drawback of handcraft feature engineering, we established novel machine learning models to automatically learn the characteristics of PAC features obtained and classify them to identify SOZ. Besides, to conquer the imbalance of datasets, we introduced novel feature-wise/class-wise re-weighting strategies in conjunction with classifiers. In addition, we proposed the time-series nest cross-validation to provide more accurate and unbiased evaluations for this model. Seven patients with focal cortical dysplasia were included in this study. The experiment results not only illustrate that the significant coupling at band pairs of slow waves and HFOs exists in the SOZ when compared with the NSOZ but also indicate the effectiveness of PAC features and the proposed models with better classification performance.

## 1. Introduction

Epilepsy is a common neurological disease characterized by epileptic seizures, which affecting 50–70 million people globally [1, 2, 3]. Approximately two-thirds of epileptic patients find that their symptoms are adequately controlled by antiepileptic drugs, while the remaining about one-third fail with medication treatment [4, 5, 6]. For the patients who have medically refractory epilepsy, epilepsy resection surgery may be the only possible effective option [7]. Since the aim of epilepsy resection surgery is the resection of the epileptogenic focus, it is crucial to localize as accurately as possible the brain region where the seizures originating from—termed as the seizure onset zone (SOZ), in the pre-surgical evaluation [8].

Electroencephalography (EEG) has been considered as one of the most important techniques in the diagnosis and treatment of patients with epilepsy during the pre-surgical process [9, 10, 11, 12]. In particular, electrocorticogram (ECoG) plays a significant role in the delineation and localization of SOZ in the pre-surgical treatment due to the high precision [13, 14]. In addition, as a major recording method in the clinical diagnosis of epilepsy, interictal ECoG is widely used for epileptic focus detection [15, 16]. In clinical practice, seizure onset zone detection is generally judgment by epileptologists visually examining and manually capturing the character of seizures from long-term ECoG recordings. However, the long-term visual inspection of ECoG is time-consuming, laborious, and error-prone [17, 18]. Therefore, it is necessary to establish advanced feature extraction and automatic SOZ localization methodologies to supply technical support in the diagnosis and treatment of epilepsy.

The interaction between different frequency bands of brain rhythms has attracted much attention from investigators particularly for the purpose of SOZ localization over the last ten years. Phase-amplitude coupling (PAC), as one typical form of brain rhythm interaction, has been considered as a promising biomarker for SOZ identification in recent investigations. The study conducted by Edakawa et al. [19] have investigated that PAC is more useful to detect ictal state than by employing high *γ* amplitude alone. Ibrahim et al. [20] have found that PAC between high frequency oscillations (HFOs) and alpha band is strong in SOZ when compared with non-epileptic zones. Surveys such as that conducted by Amiri et al. [21] have investigated that PAC between high-frequency rhythms (gamma/ripple) and slow waves is more evident in SOZ compared to normal regions. In another study, Cámpora et al. [22] has revealed that higher PAC exists in SOZ than the brain region far from the SOZ. Besides, Ma et al. [23] have illustrated that PAC of intracranial EEG during inter-and pre-seizure stage can provide an accurate reference for epileptogenic zone localization. In addition, Elahian et al. [24] has revealed that the logistic regression classifier based on PAC between the phase of 4–30 Hz and high gamma amplitude can effectively identify SOZ. Moreover, the study from Varatharajah et al. [25] has reported the feasibility to combine PAC and support vector machine (SVM) for the automate SOZ identification. Nevertheless, there is a lack of comprehensive consideration for PAC of interictal ECoG among wider frequency bands especially in the fast ripple band, which is one emphasis for our research. Most importantly, given that the PAC features may vary among different individuals and change with time, it is necessary to conduct automatic feature extraction and classification approaches for SOZ identification. Over the past few years, classical machine learning algorithms and artificial neural networks play a crucial role in disease diagnosis because of their automatic learning ability from local patterns of inputs. However, there is little literature on automatic SOZ identification under the imbalanced situation. This is the other issue addressed in our article.

Therefore, in this paper, we analyzed the characteristics of PAC of interictal ECoG recorded from seven patients (adult: four, child: three) based on the mean vector length modulation index (MVL-MI) method and statistical measures including mean and Mann-Whitney U test, where the PAC features were calculated by the sub-bands signals of ECoG between 0.5–560Hz. Besides, we established machine learning models including SVMs, light gradient boosting machine (LightGBM), and 2-D convolutional neural network (CNN) in conjunction with re-weighting technologies and proposed time series nest cross-validation (TSNCV) based on PAC features for the SOZ identification in the imbalanced input case. We therefore hypothesis that: (1) There exists a strong coupling between fast ripple and slow waves in SOZ for adult patients. (2) The coupling is not obvious for child patients when compared with adult patients; (3) The proposed models have a better prediction for SOZ identification for adult patients. The main contributions of this study can be briefly summarized as follows: (1) We comprehensively analyzed the PAC features not only between ripple and slow waves but also between fast ripple and slow waves. (2) We observed the difference in PAC between adult patients and child patients. (3) We employed five machine learning models for SOZ prediction. (4) We converted the PAC features among different band pairs to image for the first time. (5) We proposed a 2-D CNN model based on the image of PAC features for the SOZ localization. (6) Given the imbalance between the number of SOZ and NSOZ, we weighted the hyperparameters in SVMs and introduced the focal loss function and class-weighted focal loss function in the proposed 2-D CNN model, respectively. (7) In view of PAC features that may vary change over time, we designed a TSNCV for the prediction of SOZ more accurately and unbiased.

## 2. Materials and Methods

It is demonstrated in Fig. 1(A), the main work of the paper consists of three parts. The first is to analyze ECoG recorded from patients using PAC approach. Secondly, PAC features are analyzed using statistical measures. The last one is to classify the PAC features by employing the proposed models.

**Figure 1:**
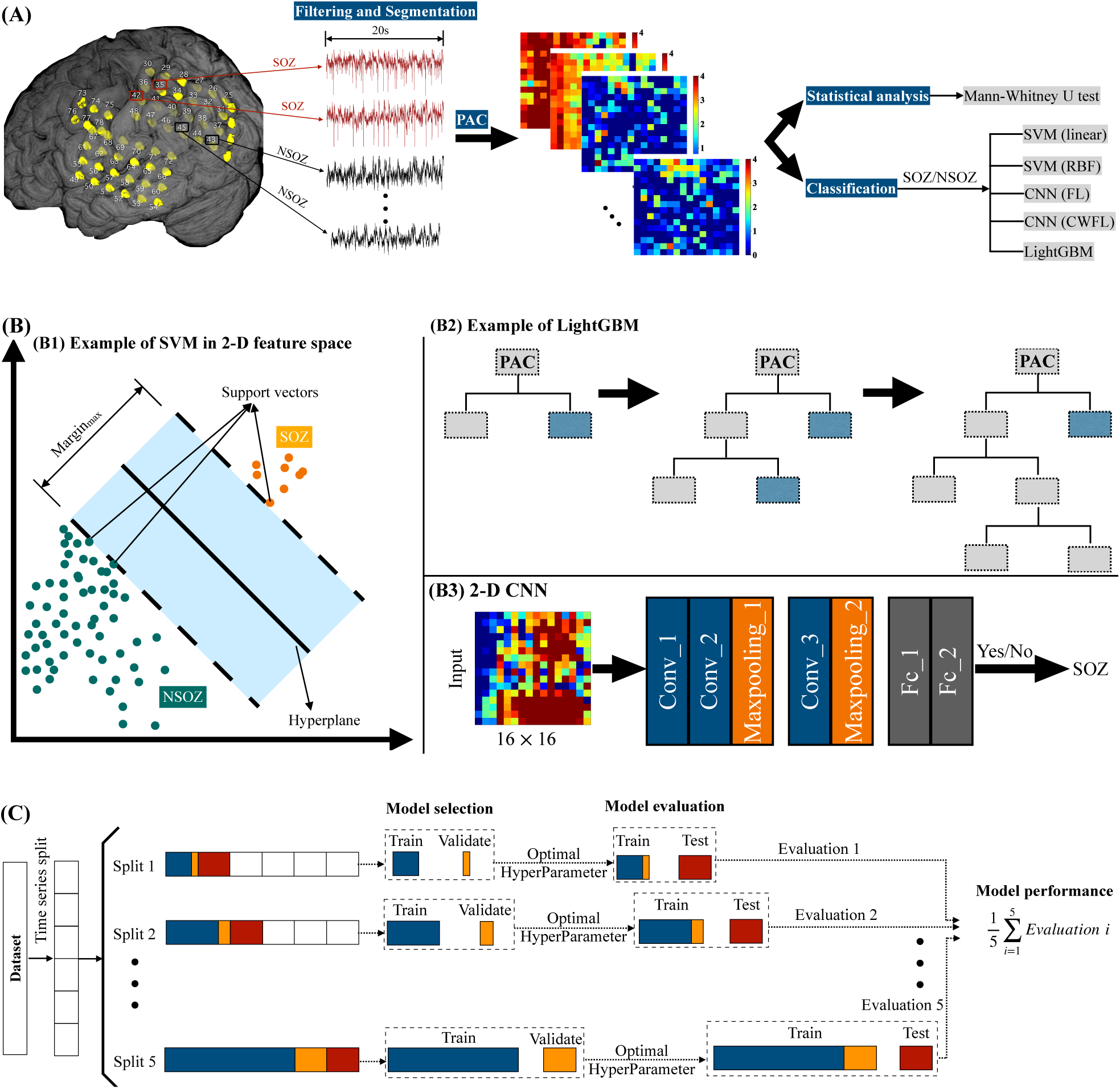
The scheme of PAC classification, the three classification models, and the proposed TSNCV. (A) The work scheme contains ECoG recording, PAC calculation and display by comodulogram, statistical analysis based on comodulogram, and classification using five models. (B) Example of SVM in 2-D feature space, Example of LightGBM, and the established 2-D CNN. (B1) is the example of SVM for simple binary classification in 2-D feature space. (B2) is the example of leaf-wise tree growth for LightGBM. (B3) is the architecture of the established 2-D CNN model. The input is the comodulogram with the size of 16 × 16 that analyzed by PAC method. There are three convolutional layers and two fully connected layers named as *Conv*_1, *Conv*_2, and *Conv*_3, *Fc*_1, and *Fc*_2, respectively. MaxPooling is respectively followed by *Conv*_2 and *Conv*_3. (C) The working principle of TSNCV. The dataset is firstly chronologically split into 5 splits. For each splits, there are training subset (denoted by dark blue color), validation set (denoted by dark orange color), and test set (denoted by brown color), in which the training subset and validation set are denoted by selecting the former 80% and the latter 20% of the training set. Firstly, the training subset is used for training while validation set is used for validation in conjunction with grid search to select the optimal hyperparameters. Hereafter, the training subset and validation set are used as training sets with the selected optimal hyperparameters, while the test set is used for testing to supply an evaluation of model performance. The final evaluation is by taking the average value of evaluations for all five splits.

### 2.1. Patients

We included the continuous one hour of interictal electrocorticogram (ECoG) data from seven patients (male: 5, female: 2; age: 5–39 years) with focal cortical dysplasia (FCD). All seven patients fell into FCD type 2A/FCD type 2B and were with the outcome of seizure free after surgery which assessed by the Engel classification at the last follow-up clinic visit [26]. The data were acquired at Juntendo University Hospital in Japan from November 2016 to July 2018. The location of electrodes was determined based on a non-invasive diagnostic protocol including seizure semiological evaluation, interictal scalp electroencephalogram (EEG), magnetic resonance imaging (MRI), fluorodeoxyglucose-positron emission tomography, psychomotor-development testing, and video-EEG monitoring for each patient before surgery [27]. The research was approved by the ethics committee at Juntendo University Hospital and the Tokyo University of Agriculture and Technology, and all patients signed consent forms.

### 2.2. ECoG Acquisition

The ECoG signals were recorded using the Neuro Fax digital video EEG system (Nihon Kohden, Inc., Tokyo, Japan) with a sampling frequency of 2,000 Hz. Epileptologist verified the number of electrodes for each patient based on the observations from ECoG recording. The range of the number of electrodes for seven patients was 36–76. The SOZ and NSOZ were labeled to corresponding electrodes according to the judgment from two epileptologists. The information of number of electrodes labeled by SOZ, number of electrodes labeled by NSOZ, lesion site, location, pathology, and duration of follow-up for seven patients were summarized in Table 1 [28, 29].

**Table 1:** Summary of information for seven patients.

| Patient ID | #SOZ electrodes | #NSOZ electrodes | Lesion Site | Location | Pathology | Duration of follow-up |
| --- | --- | --- | --- | --- | --- | --- |
| Pt1 | 7 | 63 | <sup>1</sup>Lt superior parietal lobule | Cortical surface | Type 2B | 5 years |
| Pt2 | 10 | 56 | Lt superior parietal lobule | Bottom of sulcus | Type 2B | 5 years |
| Pt3 | 16 | 60 | Lt angular gyrus | Bottom of sulcus | Type 2A | 5 years |
| Pt4 | 10 | 40 | Lt dorsal superior frontal gyrus | Bottom of sulcus | Type 2B | 3 years |
| Pt5 | 3 | 33 | <sup>2</sup>Rt dorsal middle frontal gyrus | Cortical surface | Type 2B | 3.5 years |
| Pt6 | 3 | 57 | Lt dorsal superior temporal gyrus | Cortical surface | Type 2B | 3 years |
| Pt7 | 6 | 36 | Lt cingulate gyrus | Bottom of sulcus | Type 2B | 3.5 years |

<sup>1</sup>Lt is the abbreviation of left.
<sup>2</sup>Rt is the abbreviation of right.

### 2.3. Preprocessing

Continuous one-hour interictal ECoG data were band-pass filtered by employing finite impulse response with Kaiser window (scipy.signal.firwin function, Python), in which the length of filter was set to odd in order to avoid zero response at the Nyquist frequency [30]. For each electrode, the ECoG data were filtered between 0.5 and 24 Hz with 0.5 Hz/1 Hz/2 Hz bandwidth for the low-frequency rhythms and 80–560 Hz with a bandwidth of 30 Hz for the high-frequency components. Specifically, in order to analyze the coupling of low-frequency signals in detail, especially for the *δ* band (0.5–4 Hz) and *θ* band (4–7 Hz), different bandwidths were set, in which the bandwidth of frequency band for 0.5–1 Hz, 1–8 Hz, and 8–24 Hz were set to 0.5 Hz, 1Hz, and 2 Hz, respectively. Then, the instantaneous phase of low-frequency filtered data and the instantaneous amplitude of high-frequency filtered signals were calculated by using Hilbert transform method [31]. Then each hour-long section of phase/amplitude data were fragmented into 180 segments, each of 20-s duration.

### 2.4. Calculation of Mean Vector Length Modulation Index

The PAC value was quantified employing MVL-MI measure [32]. Specifically, the low-frequency phase and high-frequency amplitude are denoted as *A*_*amp*_(*t*) and *ϕ*_*pha*_(*t*), respectively. The *A*_*amp*_(*t*) is projected on *ϕ*_*pha*_(*t*) to form to a complex component as 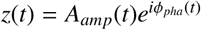. The raw modulation index is then defined as 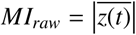 by calculating the absolute of mean vector of *z*(*t*), the length of *z*(*t*). From the definition of *z*(*t*), the length of *z*(*t*) is based on the amplitude. So the surrogate data approach is used to z-scored the *MI*_*raw*_ by using the mean, standard deviation of 100 surrogate data, where the mean and standard deviation of surrogate data are denoted as *μ* and *σ*, respectively. The final modulation index is defined as *MI* = (*MI*_*raw*_ − *μ*)/*σ*. It is noted that the surrogate data is generated under the null hypothesis of no modulation by randomly shuffling the amplitude time series to disrupt the phase-amplitude relationship [33, 34].

For multiple sub-bands PAC calculation, phase-amplitude comodulogram is introduced to graphical display the PAC values over a grid of frequencies [35]. The horizontal axis is used to represent the frequency of phase and the ordinate axis is applied to display the frequency of amplitude, as well as the PAC value at the corresponding band pair is denoted by using a pseudocolor plot with hot color representing high value.

### 2.5. Classifiers

In this paper, both classical machine learning algorithms and artificial neural networks were employed for SOZ identification based on PAC features. In detail, the five models including SVM with linear kernel, SVM with radial basis function (RBF) kernel, LightGBM, 2-D CNN with focal loss function, and 2-D CNN with class-weighted focal loss function were introduced. Besides, the input for 2-D CNN is the comodulogram with the size of 16 × 16 for each segment, and for SVM and LightGBM, the input is expected to be flattened the feature dimension from the 2-D array with the size of 16 × 16 to a 1-D array with the size of 256.

#### 2.5.1. Support Vector Machine Model

SVM is a common supervised learning model for real-world binary classification, which is based on statistical learning theory [36]. The motivation of SVM is to project nonlinear feature vectors onto a high-dimensional feature space by using kernel functions and to determine the optimal separating hyperplane in the feature space to most effectively separate the two-class samples [37]. A simple example of SVM in 2-D feature space is as illustrated in Fig. 1(B1). The optimal separating hyperplane is defined by which not only classifying the two categories but also ensuring that the margin between the samples from two categories and the hyperplane segmentation boundary is max. Common kernel functions include linear kernel, sigmoidal kernel, polynomial kernel, and RBF kernel. In this study, the SVM with linear kernel and the SVM with RBF kernel were employed.

There are two typical hyperparameters *C* and *gamma* in SVM, in which the hyperparameter *C* in SVM is used to determine the penalty for misclassifying during the fit of the model and the hyperparameter *gamma* is employed to provide curvature weight of the decision boundary. Since the ratio of two-class samples is imbalanced as listed in Table 1, the method to handle imbalanced classification for SVM in this study is to weight the hyperparameter *C* in proportion to the importance of the two classes. Specifically, decreasing the weight to the majority class while increasing the weight for the minority class to prevent misclassification. As is shown in formula below.

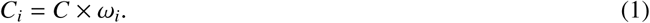

Where *C* is the penalty, *C*_*i*_ is the *C*value of class *i, ω*_*i*_ is a weight inversely proportional to class *i*.

#### 2.5.2. Light Gradient Boosting Machine Model

LightGBM is a novel distributed gradient boosting model based on decision tree algorithm [38], as presented in Fig. 1(B2). Besides, LightGBM grows trees leaf-wise and implements histogram-based algorithms, in which the leaf-wise tree growth algorithm selects the leaf with the highest loss decrease and the histogram-based algorithms arranges continuous features to discrete bins to accelerate and memory-saving [39]. In this study, the LightGBM Python module is used [40]. The weight for each class is automatically adjusted by setting the parameter *class*_*w*_*eight* to *balanced* to handle the imbalanced classification issue. In addition, we also conduct parameters tuning to improve the model performance, in which the parameters consist of the number of leaves per tree *num_leaves*, the maximum tree depth *max_depth*, the boosting learning rate *learning_rate*, and the minimal number of data in one leaf *min_data_in_lea f*.

#### 2.5.3. 2-D Convolutional Neural Network Model

Since each segment of phase-amplitude comodulogram can be converted into an image with 16 × 16 pixels. In this study, we proposed a 2-D CNN network structure for focal identification based on comodulogram results obtained. As described in Fig. 1(B3), two convolution blocks are included in the proposed model. The first convolution block consists of two convolutional layers, a batch normalization after each convolutional layer, and a MaxPooling layer. The former and the latter convolutional layers have 32 and 64 convolution kernels, respectively. The kernel size is set to 3 × 3 with the stride of 1 in both the two convolutional layers. Following is the MaxPooling layer with a size of 2 × 2. The second convolution block comprises one convolutional layer of 128 kernels with a kernel size of 3 × 3 and a stride of 1, a batch normalization, and a MaxPooling layer of 2 × 2. The rectified linear unit (ReLU) activation function and the He Uniform initialization are used in all three convolutional layers. Stacked behind the two convolution blocks are two fully connected layers. The first fully connected layer uses the ReLU activation and the dropout rate of 0.5, while the second one applies a sigmoid activation. The values of epoch and batch size are set to 100 and 128, respectively. Besides, early stopping is used in this model to prevent overfitting. The proposed model is implemented in Keras 2.3.1 with Tensorflow 1.14.0 backend [41]. The model run in the server configured with one NVIDIA Tesla 418 Driver.

#### 2.5.4. Focal Loss Function

As can be seen in Table 1, the comodulogram results used for classification are imbalanced, in which the ratio of SOZ samples to NSOZ samples for patients is from 1 : 3.75 to 1 : 19. To address the imbalanced classification, the focal loss function is introduced in this work. Focal loss function is a modulating term of the cross-entropy (CE) loss function, which makes the model focus training on hard samples by down-weighting of natural samples, and the definition is as follows [42].

Taking the binary classification in the cross entropy loss as an example, the classification loss is the summation of the entropy of each training sample where the weight of each sample is the same. The formula is shown as

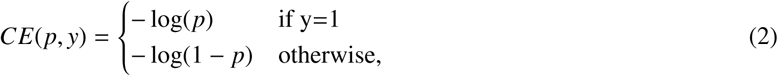

where *y* is the label of two classes in the binary classification model with the range of *y* ∈ {±1}, and *p* ∈ {0, 1} is the model predicted probabilities of the samples with label *y* = 1. For simplicity, defining *p*_*t*_ as

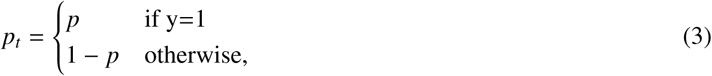

and *CE*(*p; y*) can be updated to

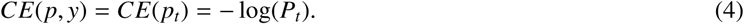

To deal with the obstacle that the number of samples for two classes varied greatly, a common practice is to introduce the weights to both the positive samples and negative samples. Down-weighting the negative samples if the negative class frequency is higher, while increasing the weights of positive samples with lower positive class frequency. So the weighting factor *α* ∈ {0, 1} is introduced to control the shared weights of the two-class samples to the total loss. In a similar way to define *p*_*t*_, defining *α*_*t*_ as

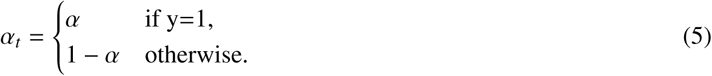

And the balanced cross entropy can be defined as

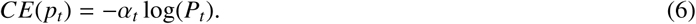

Moreover, to control the weights of hard samples and easy samples, a modulating factor (1 − *P*_*t*_)^*γ*^ is introduced to focus learning on hard samples by reducing the weights of easy samples. So the *α*-balanced variant of the focal loss is defined as

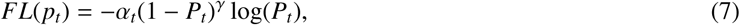

where *γ* is focusing parameter with the range of *γ* ≥ 0. In this study, the ranges of parameters *α, γ* were set to [0.75, 0.9, 0.99, 0.999, 0.9999] and [0.5, 1, 2, 5], respectively.

#### 2.5.5. Class-Weighted Focal Loss Function

The other loss function we introduced to tackle the issue of imbalanced input is the class-weighted focal loss function. It improves the performance of loss function on imbalanced data by introducing a weighting factor that is inversely proportional to the effective number of samples [43]. Specifically, *N*(*N*_≥_1) is assumed as the volume of data in the feature space of a class. The effective number of samples is defined as

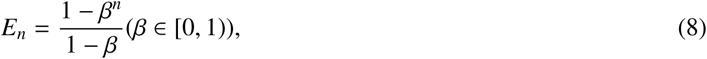

where *n* is the number of samples for the class, and 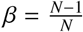. The weighting factor is simply defined as 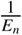 to balance the loss. So the class-weighted focal loss function is defined as

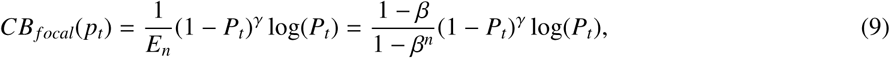

for the experiment of 2-D CNN with class-weighted focal loss function in this paper, the range of parameters *β, γ* were set to [0.75, 0.9, 0.99, 0.999, 0.9999] and [0.5, 1, 2, 5], respectively.

### 2.6. Time Series Nest Cross-Validation

To tune hyperparameters and ensure a robust evaluation of the model, a cross-validation measure is used in this work. In particular, due to the comodulogram changed over time, the time series nest CV based on our classification models is proposed. Moreover, due to the limited comodulogram dataset, the training dataset is split into a training subset and validation set according to the chronological order to prevent overfitting during model training. As presented in Fig. 1(C), the time series nest cross-validation (CV) consists of the inner loop and outer loop, in which the inner loop is normal CV with a grid search function for tuning parameters, while the outer loop is used to estimate the performance with the optimal hyperparameters obtained from the inner loop. Specifically, the dataset is chronologically split into *K* + 1 folds using sklearn.model selection.TimeSeriesSplit function [44]. In the *K*th split, the K folds are used as the training set, and the (*K* + 1)th folds as the test set. *K* is set to five in this work. Besides, the training set is further split into a training subset and validation set by selecting the former 80% and the latter 20% of the training set, respectively. For each inner loop, the model is trained on the training subset and validated on the validation set to select the optimal hyperparameters. Afterward, the model is trained on the training set with optimal hyperparameters obtained from each inner loop and estimates the model performance on the test set. The final estimation of the outer loop is obtained by calculating the average of estimations from all splits.

### 2.7. Model Evaluation

The metric used in the model evaluation is the area under the curve (AUC) of receiver operating characteristics. Moreover, to get the probability prediction that each channel of each patient belongs to class SOZ, we take the mean of the probability prediction for each channel of all splits and display the mean values intuitively by using a horizontal bar chart and heat map of MRI.

## 3. Results

In this section, we calculated PAC features with one hour interictal ECoG data recorded from each patient by employing the MVL-MI approach illustrated in Section 2.4. In detail, we computed the PAC values of each segment between the phase of 0.5–24 Hz with various 0.5 Hz/1 Hz/2 Hz bandwidth and the amplitude of 80–560 Hz with 30 Hz bandwidth, yielding 16 and 16 sub-bands each. The PAC values of each segment at different band pairs were displayed using a comodulogram. We firstly tested these comodulograms with statistical analysis measures including mean, violin plot, and Mann-Whitney U test. Hereafter, we conducted a classification to identify the SOZ using the established models.

### 3.1. Comodulogram with Respect to Raw ECoG

The time-domain in SOZ (brown) and NSOZ (green), as well as the PAC values with comodulograms were represented in Fig. 2 for both groups (adult vs child). It is observed that a prominent relation exists for adult patient at the band pairs of *δ/θ/α/β* − *HFO* in the SOZ compared to in the NSOZ of child patient. Similar evidence was observed in SOZ with spikes for time-domain analysis.

**Figure 2:**
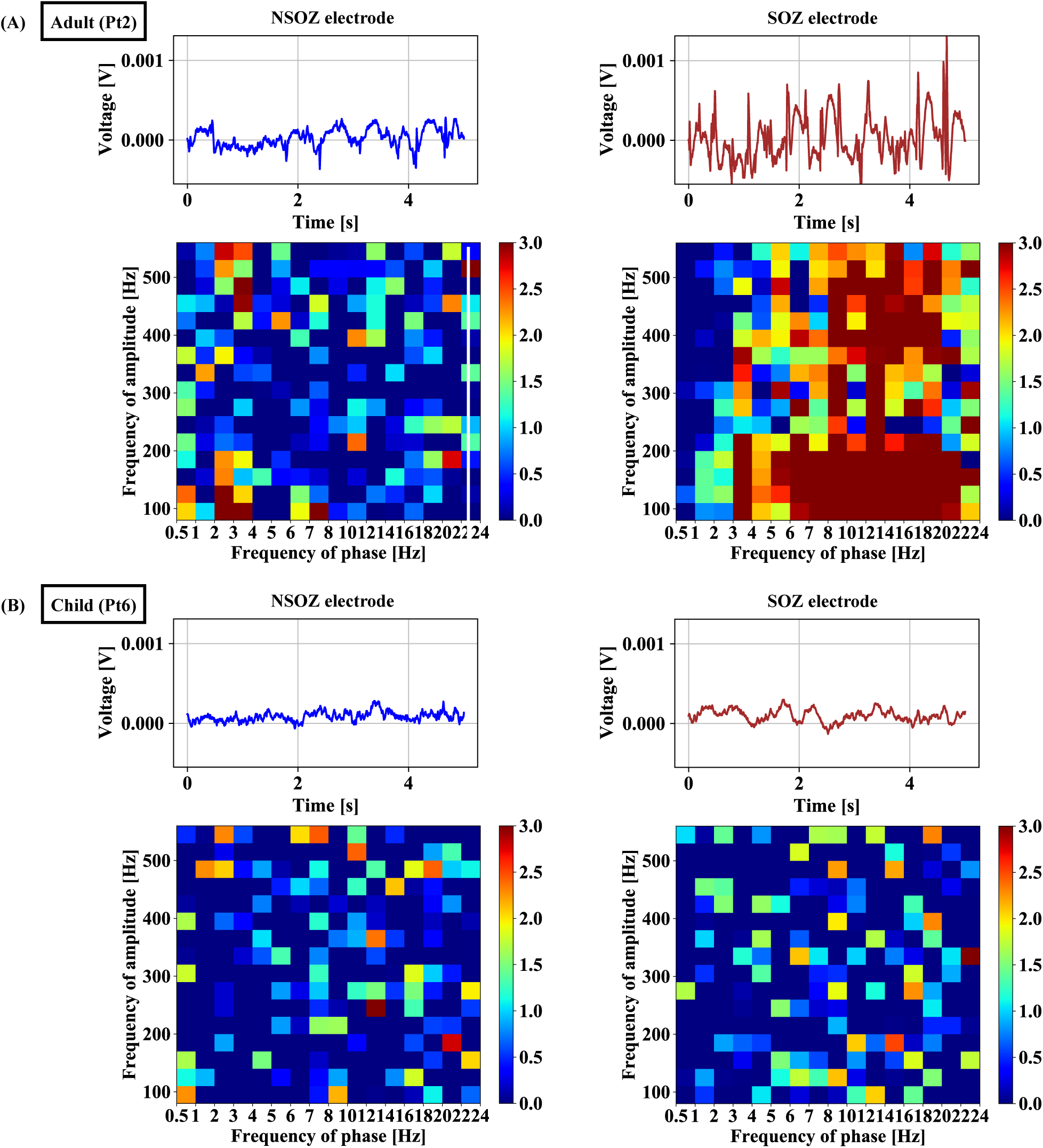
Raw interictal ECoG and comodulogram plot in SOZ and NSOZ for adult (Pt2) shown in sub-figure (A) and child (Pt6) patients displayed in sub-figure (B).

### 3.2. Statistical Analysis with Two Groups (Adult vs Child)

To intuitively observe the differences of coupling between SOZ and NSOZ for each patient, we took the mean comodulogram of both SOZ group and NSOZ group. The number of samples of SOZ and NSOZ for each patient was listed in Table 1. As seen in Fig. 3(A) and Fig. 4(A), it was shown that there exist apparent differences in mean comodulogram between SOZ and NSOZ. Besides, it could be visually observed that the coupling for SOZ mainly happened at band pairs of *δ/θ/α* − *HFO* when compared with NSOZ for four adult patients. While for three child patients, the coupling phenomenon is various. Specifically, the differences of coupling between SOZ group and NSOZ group are weak (Pt5) and even the coupling value for SOZ is lower when compared to NSOZ (Pt7). While there exists obvious coupling in the band pair of *δ/θ/α* − *ripple* and *δ/θ* − *f astripple* for Pt6. Furthermore, the total coupling strength for four adult patients is much stronger than PAC strength for three child patients.

**Figure 3:**
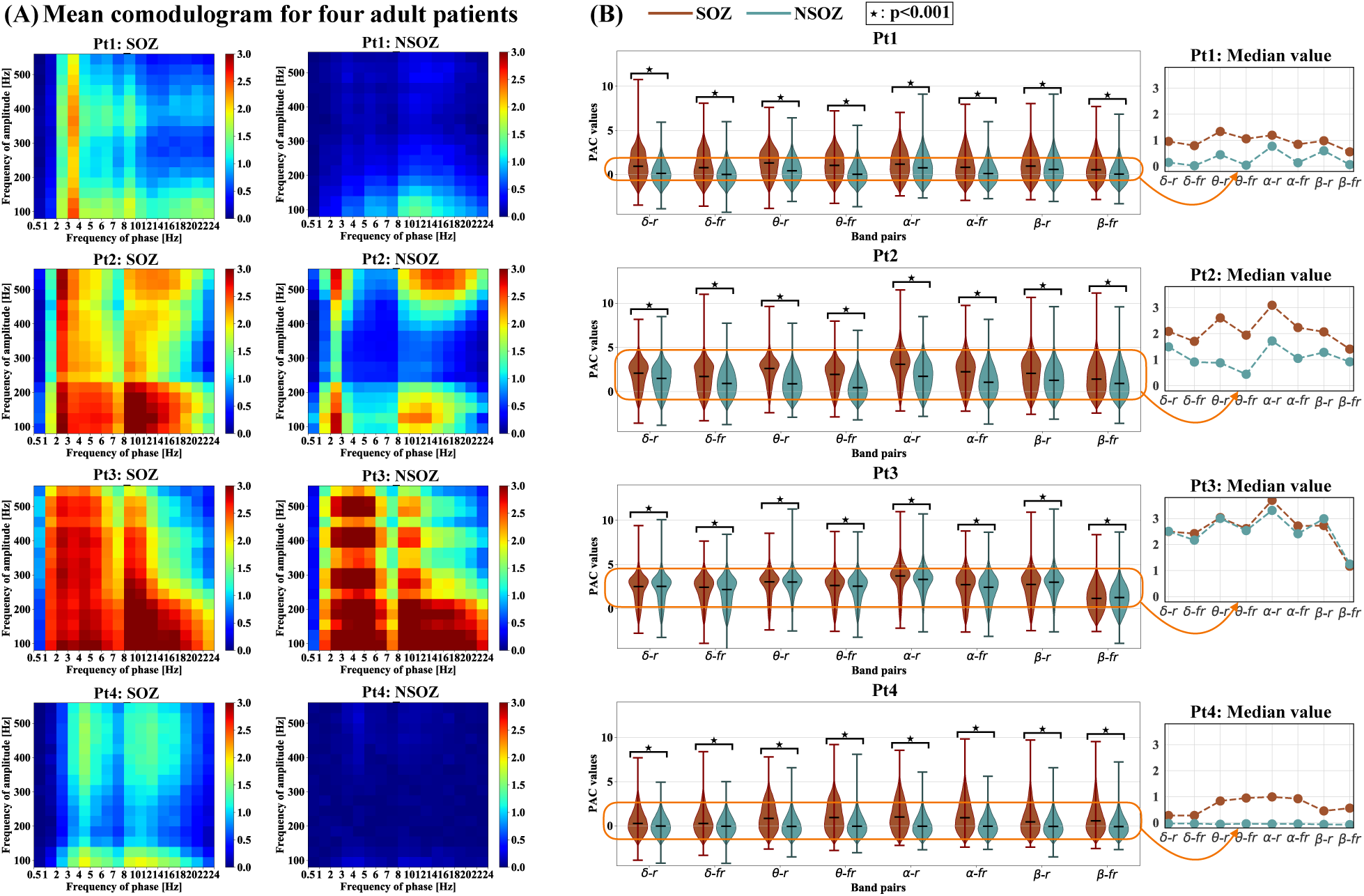
The mean comodulogram and the Mann-Whitney U test results at eight band pairs in both SOZ and NSOZ for four adult patients. (A) The mean comodulogram of SOZ and NSOZ for each adult patient. For each mean comodulogram, x-axis denotes the frequency of phase, the y-axis indicates the frequency of amplitude, as well as the pseudocolor represented the PAC value at different band pairs. (B) The Mann-Whitney U test results of SOZ and NSOZ for four adult patients at eight band pairs. The median value of each band pair for both SOZ and NSOZ is displayed zoom in the most right sub-figure. The eight band pairs are *δ* − *r, δ* − *f r, θ* − *r, θ* − *f r, α* − *r, α* − *f r, β* − *r*, and *β* − *f r*, representing *δ* − *ripple, δ* − *f astripple, θ* − *ripple, θ* − *f astripple, α* − *ripple, α* − *f astripple, β* − *ripple*, and *β* − *f astripple*, respectively, where the frequency range of *δ, θ, α, β, ripple*, and *f astripple* is 0.5–4 Hz, 4–8 Hz, 8–12 Hz, 12–24 Hz, 80–250 Hz, and 250–560 Hz, respectively. X-axis and y-axis imply types of band pairs and PAC values, respectively. Brown color implies the SOZ while green color indicates the NSOZ. ⋆indicates *p* < 0.001.

**Figure 4:**
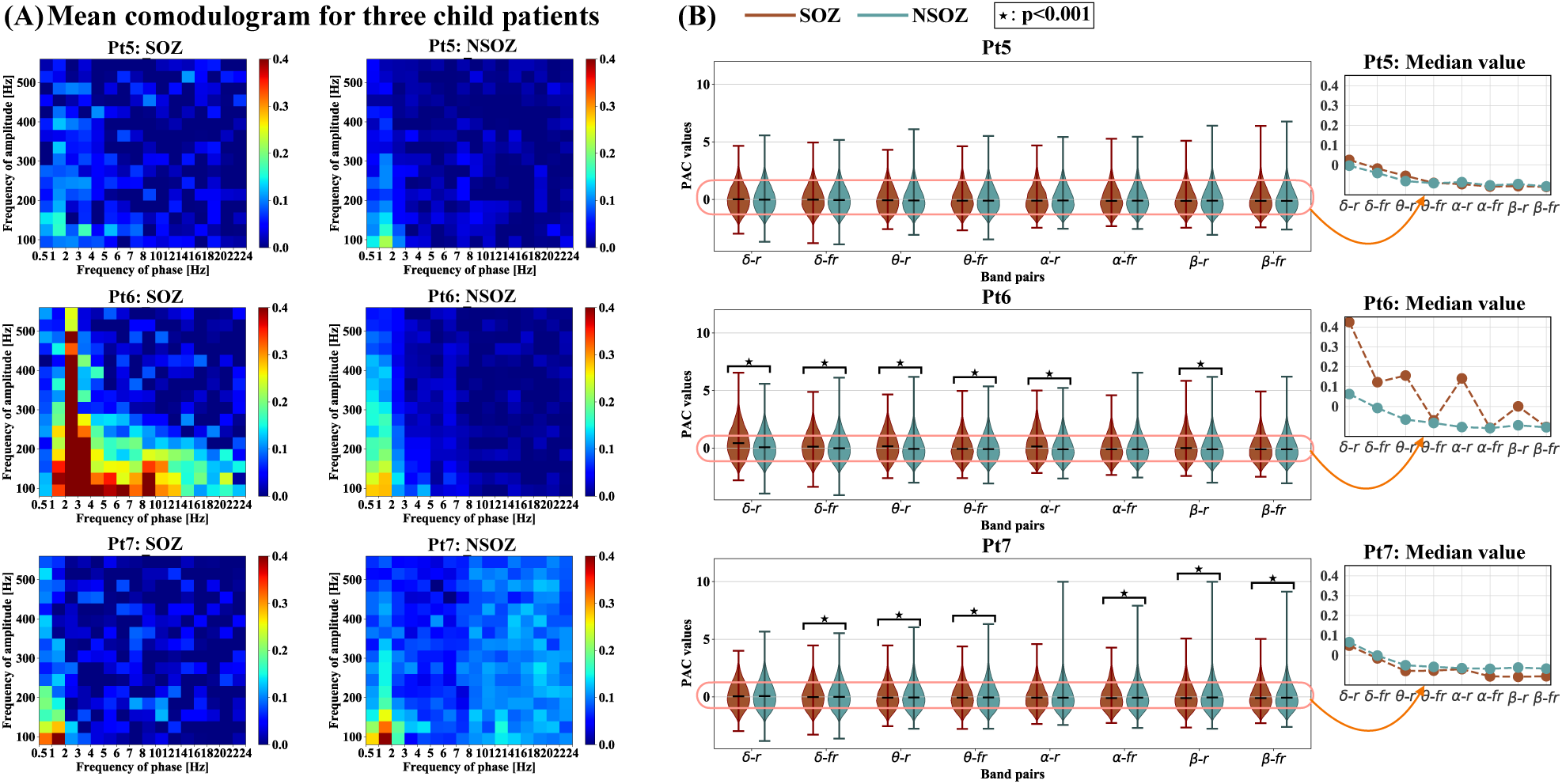
The mean comodulogram and the Mann-Whitney U test results at eight band pairs in both SOZ and NSOZ for three child patients, shown in sub-figure (A) and sub-figure (B), respectively.

As illustrated above, the coupling mainly focuses on the band pairs of *δ/θ/α/β*−*HFO*, therefore, the PAC values of each electrode with seven patients were extracted for statistical analysis. Fig. 3 (B) and Fig. 4 (B) show the violin plot approach to show the statistical differences of coupling properties with the combinations of eight bands (*δ* − *ripple, δ* − *fastripple, θ* − *ripple, θ* − *fastripple, α* − *ripple, α* − *fastripple, β* − *ripple, β* − *fastripple*). In that case, the SOZ and NSOZ group for each patient were used for analysis. The frequency ranges of each band for analysis were represented as *δ* (0.5–4 Hz), *θ* (4–7 Hz), *α* (8–12 Hz), *β* (13–24 Hz), *ripple* (80–250 Hz) and *fastripple* (250– 560 Hz), respectively. It is demonstrated that the PAC values for adult patients are much higher than those for child patients. Moreover, there exist individual differences in PAC values among different individual patients.

The Mann-Whitney U test was employed to test the statistically significant difference in PAC values distributions between SOZ and NSOZ for seven patients. Specifically, PAC values of SOZ and NSOZ at eight band pairs for each patient were extracted. Every band pair of PAC values distribution was displayed by violin plot for both SOZ and NSOZ. Mann-Whitney U test was introduced to test whether there was a significant difference in the PAC values distribution between the SOZ and NSOZ at the eight designated band pairs at the significance level of 0.001. As shown in Fig. 3(B), the median PAC value of SOZ was higher than that of NSOZ at the band pair of *δ* − *fastripple, θ* − *ripple, θ* − *fastripple, α* − *ripple*, and *α* − *fastripple* for all four adult patients, which is consistence with the observation in Fig. 3(A). Besides, the results of the Mann-Whitney U test showed the statistically significant difference between the PAC values of SOZ and NSOZ for adult patients at all eight band pairs at a significant level of 0.001. While in the case of child patients (Pt5), there is no significance between SOZ and NSOZ, as observed in Fig. 4(B). Moreover, the Mann-Whitney U test results between adult SOZ group and child SOZ group as well as results between adult NSOZ group and child NSOZ group is summarized in Fig. 5. It is illustrated that the coupling strength of adult SOZ group and adult NSOZ group are significantly stronger when compared to child SOZ group and child NSOZ group, respectively.

**Figure 5:**
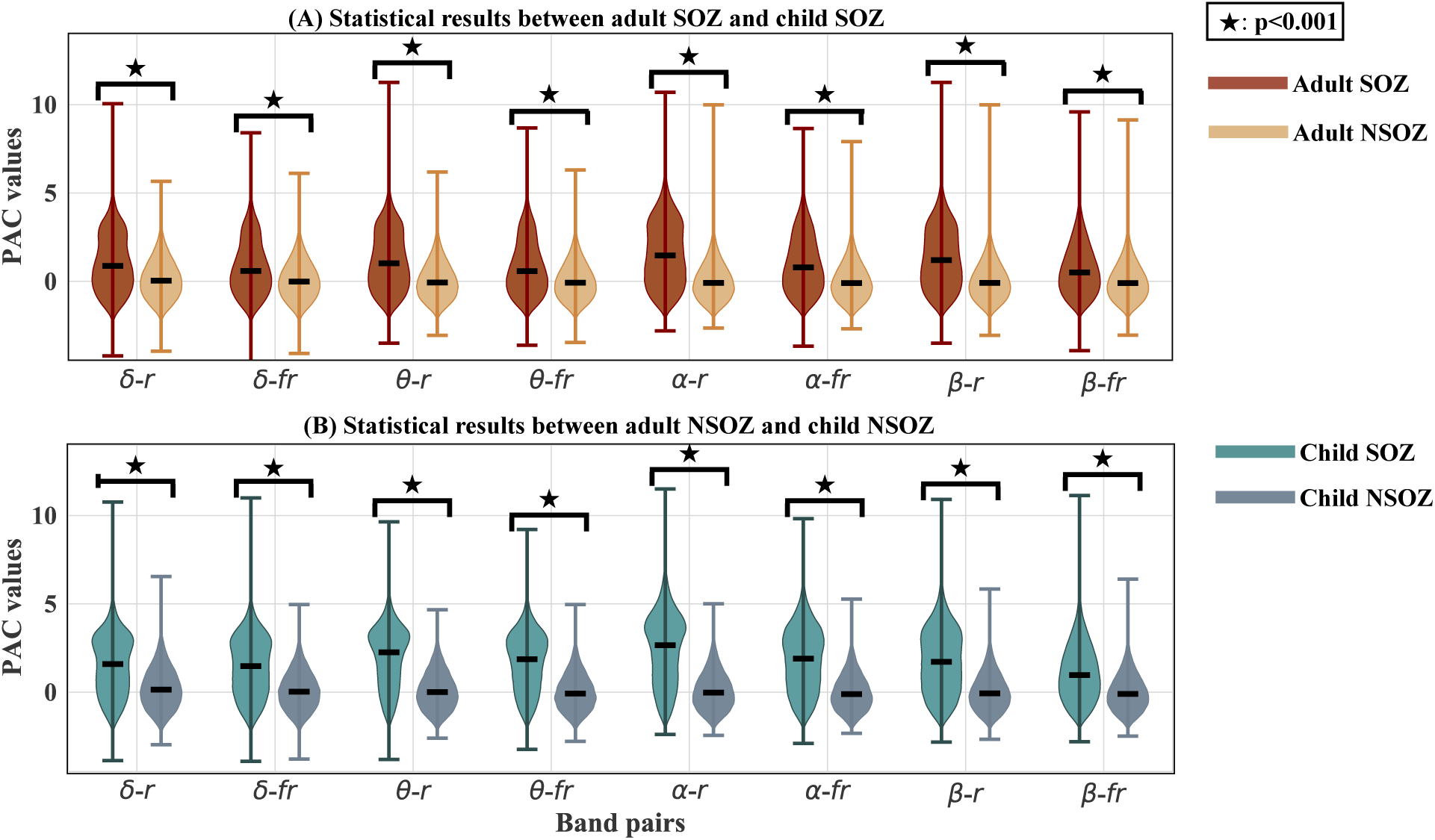
Statistical results between adult SOZ group and child SOZ group, as well as between adult NSOZ group and child NSOZ group. Brown color, yellow color, green color, slategrey color indicate the adult SOZ, child SOZ, adult NSOZ, child NSOZ, respectively. ⋆implies *p* < 0.001.

### 3.3. Results of Classification

We used the proposed SVM with linear kernel, SVM with RBF kernel, LightGBM, 2-D CNN with focal loss function, 2-D CNN with class-weighted focal loss function in conjunction with the proposed TSNCV to classify the SOZ and NSOZ based on the PAC features for the individual patient. The TSNCV was set to splut five times. The final result was presented by mean value with standard deviation. AUC was used to evaluate model classification performance. Table 2 summarized SOZ prediction results using five classifiers for seven patients. It is demonstrated that the classifiers can better detect the SOZ electrodes of adult patients based on the PAC features, while it fails to recognize the SOZ electrodes of children. Besides, when compared the results of another four models (SVM with linear kernel, LightGBM, 2-D CNN with focal loss function, and 2-D CNN with class-weighted focal loss function), we observed that the results computed by SVM with RBF kernel were slightly better with the average AUC for four adult patients of 0.915. In particular, the AUC of Pt2 using SVM with RBF kernel was up to 0.963. Moreover, the AUCs of three adult patients (Pt1, Pt2, and Pt3) were higher than 0.9 computed by five classifiers.

**Table 2:** Results of AUC for seven patients using five classifiers.

| Patient | Adult Pt1 | Adult Pt2 | Adult Pt3 | Adult Pt4 | Child Pt5 | Child Pt6 | Child Pt7 | Mean of Adult | Mean of Child | Mean of all |
| --- | --- | --- | --- | --- | --- | --- | --- | --- | --- | --- |
| <sup>1</sup>SVM<sub>Linear</sub> | 0.909 | 0.940 | 0.900 | 0.847 | 0.489 | 0.780 | 0.501 | 0.899 | 0.590 | 0.767 |
| <sup>2</sup>SVM<sub>RBF</sub> | 0.912 | 0.963 | 0.938 | 0.847 | 0.493 | 0.772 | 0.492 | 0.915 | 0.586 | 0.774 |
| LightGBM | 0.904 | 0.957 | 0.935 | 0.837 | 0.505 | 0.747 | 0.494 | 0.908 | 0.582 | 0.768 |
| <sup>3</sup>CNN<sub>FL</sub> | 0.922 | 0.935 | 0.931 | 0.819 | 0.508 | 0.732 | 0.493 | 0.902 | 0.578 | 0.763 |
| <sup>4</sup>CNN<sub>CWFL</sub> | 0.909 | 0.935 | 0.902 | 0.824 | 0.509 | 0.639 | 0.503 | 0.893 | 0.550 | 0.746 |

<sup>1</sup>SVM<sub>Linear</sub> is the SVM with the linear kernel.
<sup>2</sup>SVM<sub>RBF</sub> is the SVM with the RBF kernel.
<sup>3</sup>CNN<sub>FL</sub> is the CNN with the focal loss function.
<sup>4</sup>CNN<sub>CWFL</sub> is the CNN with the class-weighted focal loss function.

Furthermore, we calculated the mean probability prediction of five splits for each corresponding electrode for four adult patients to demonstrate the probability which the electrode was classified to be SOZ based on the SVM with RBF kernel. Fig. 6(A) demonstrates that the probability prediction for SOZ electrodes was high, six out of seven for electrodes A9–A12, A32, and A37. This was much higher than for Pt1’s NSOZ electrodes. Likewise, for Pt2, Pt3, and Pt4, the probability prediction of nine (A15, A16, A29, A30, A35, A36, A41, A42, A48) out of ten SOZ electrodes (A15–A17, A29, A30, A35, A36, A41, A42, A48), 16 (A1–A16) out of 16 SOZ electrodes (A1–A16), as well as five (A9, A13, A14, A17, A18) out of ten SOZ electrodes (A9, A10, A13, A14, A17–A19, A26, A32, A38), were higher than that of corresponding NSOZ electrodes, respectively. Moreover, the location information of electrodes and corresponding values of prediction probability were demonstrated in Fig. 6(B). It was noted that the location information of electrodes A1–A16 for Pt1 and electrodes A1-A24 for Pt2 were not shown in MRI because these electrodes were placed in the cortical sulcus. In detail, for Pt1, NSOZ electrode A36 with higher probability prediction was located near the SOZ electrode A37. Besides, NSOZ electrode A11 with higher probability prediction was located next to the SOZ electrode A9 for Pt4.

**Figure 6:**
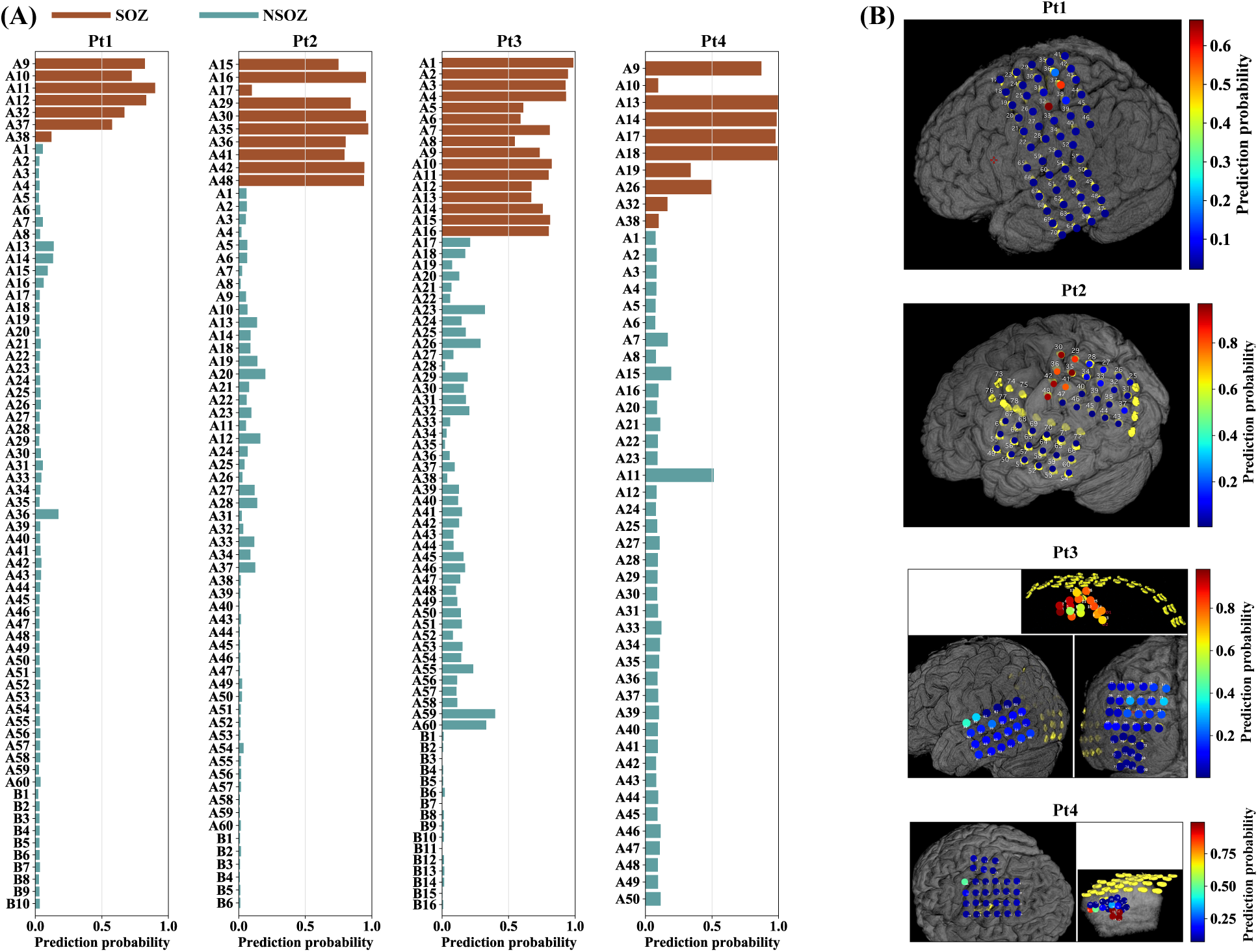
The prediction probability in bar-plot and MRI for four adult patients. (A) The bar-plot of prediction probability of each electrode for four adult patients. X-axis and y-axis denote the value of prediction probability and channel name, respectively. The brown color indicates the SOZ electrodes, while green represents the NSOZ electrodes. (B) The MRI of prediction probability for four adult patients. The pseudo color denotes the value of the prediction probability of each electrode. It is noted that the number 1–60 and 61–76 in (B) represent A1–A60 and B1–B16 in (A), respectively.

## 4. Discussion and Conclusion

Recent studies have hypothesized that PAC can reflect information transfer within neurons, especially can strengthen the synchronization between high-frequency components and specific phase of slow wave [45, 21, 33]. We analyzed the coupling characteristics of one hour interictal ECoG recorded from seven epileptic patients by employing the MVL-MI method. Since a considerable amount of literature has been published that HFO is believed a promising biomarker of epileptogenic zone (EZ) [46, 47, 48], besides, there is almost no literature to investigate PAC between fast ripple and slow rhythms for SOZ localization, we calculated PAC between low-frequency phase and the amplitude of HFOs (ripple and fast ripple) for interictal ECoG. The statistical results shown in Fig. 3 verified our hypothesis that there exists significant coupling in the SOZ at band pairs of slow rhythms and HFO when compared with NSOZ for adult patients. It might indicate the hyperpolarization of cortical neurons due to low-frequency waves co-occurring with interictal HFOs [49]. In addition, results of the mean comodulogram also demonstrated differences in specific coupling band pairs due to the individual differences. Besides, related study investigated the higher PAC values of *β* − high *γ* in ictal state were located around or within SOZ [19]. Combined our study, it implies higher PAC values both in interictal state and ictal state may be used as a promising biomarker to identify SOZ. These findings motivated PAC study in identification of SOZ.

Moreover, we firstly illustrated the differences in PAC values between adult epileptic patients and child epileptic patients presented in Fig. 3, Fig. 4, and Fig. 5. The range of PAC and the band pairs in which coupling occurs for adult patients are different from that for child patients. Compared with PAC results for child patients, the range of PAC for adult patients is much higher. It implies that coupling properties may be related to the age of the subject. Furthermore, the PAC between SOZ and NSOZ for child patients presents no significance at some band pairs. Fig. 2 indicates the stronger coupling may be correlated to spikes in time-domain. These observations suggest that PAC may be a candidate biomarker to localize the SOZ for adult patients who have prominent coupling, and it is necessary to combine another feature to confirm the position of the SOZ for the child patients whose seizures are more complicated compared to adult patients.

Given the PAC features were time-varying and individual differences, it was difficult for manual feature extraction. Classical machine learning algorithms SVM and lightGBM were employed to identify SOZ automatically in this paper. In addition, each segment of PAC at different band pairs which was displayed by comodulogram can be considered as an image with 16 × 16 pixels. Therefore, we also designed the 2-D CNN model explained in Section 2.5.3 to learn and identify characteristics of inputs automatically. Moreover, as seen in Table 1, the number of SOZ samples was far more than the number of NSOZ samples for each patient. In SVM the hyperparameter *C* is weighted to handle the imbalance of classes, in which *C* is in proportion to the importance of the classes, while in CNN we consequently introduced two types of the loss function (focal loss function and class-weighted focal loss function) to overcome the imbalanced classification. Both loss functions had a good performance for imbalanced inputs. Furthermore, we also proposed the TSNCV for the model to improve the accuracy and unbiasedness for the evaluation of classification performance. Classification results presented in Table 2 illustrated that the results by using SVM with RBF kernel were slightly better compared to another four models (SVM with linear kernel, LightGBM, 2-D CNN with focal loss function, and 2-D CNN with class-weighted focal loss function) for SOZ identification with the average AUC for four adult patients of 0.915. Last but not least, as indicated in Fig. 6(B), some NSOZ electrodes with higher probability prediction may be related to the reason that their implant positions were located close to the SOZ region.

There are three major limitations in present work that could be addressed in the future study. Firstly, the research was further limited by the small number of patients. With limited subjects, the results might not be possibly generalizable. In future research, we will consider more patients’ data. Another limitation of this research had to do with the singleness of features. It was shown in Table 2 that the SOZ of adult patients could be predicted accurately with a higher AUC using PAC alone, while it is not easy to identify SOZ by only applying PAC for child patients. Future research could improve on these results by combining PAC with other promising features in child patients. Finally, the research findings of this work were limited by the generality of the proposed model. The proposed model could supply better classification evaluation for given individuals while being a lack of applicability to new patients. Hence, to overcome the drawback of patient-specific models, effective methodologies are required such as transferring learning in future research [50, 51].

SOZ localization in the process of drug-resistant epilepsy patient diagnosis using interictal ECoG is challenging research. In this paper, we applied one of PAC calculation approaches called MVL-MI to calculated comprehensively PAC between slow-waves and HFO (including fast ripple). More importantly, we proposed novel models combined with SVMs, lightGBM, CNNs, and one form of cross-validation named TSNCV for automatic feature learning and SOZ prediction. Moreover, we performed statistical analysis on PAC features to assess the significance between SOZ and NSOZ at different band pairs. Final results support our hypothesis illustrated in Section 1.

## Data Availability

All data produced in the present work are contained in the manuscript.

## Author Contributions

Y.M. proposed the methods, wrote the program, analyzed the data, and wrote the manuscript. Y.I. and H.S. made contributions to the data collecting and improvement of the manuscript. K.F. contributed to the improvement of data acceleration processing and the model. T.T. contributed to the work design, improved the method, and revised the manuscript.

## Acknowledgments

This work was fully supported by JST CREST (Grant No. JPMJCR1784).

